# Risk and mitigation of aerosolisation from lung function testing: results from the AERATOR study

**DOI:** 10.1101/2021.03.06.21253033

**Authors:** S Sheikh, F Hamilton, GW Nava, F Gregson, D Arnold, C Riley, J Brown, AERATOR group, B Bzdek, J Reid, N Maskell, JW Dodd

**Author notes:** <u>Corresponding author:</u> Dr James W. Dodd MB ChB PhD FRCP, Consultant Senior Lecturer Respiratory Medicine, Academic Respiratory Unit, University of Bristol, Level 2 Learning & Research Building, Southmead Hospital, Bristol BS10 5NB, UK. These authors contributed equally. These authors contributed equally (N Maskell, JW Dodd). AERATOR group (in alphabetical order): The AERATOR group consists of (in alphabetical order): Arnold, D; Brown, J; Bzdek, B; Cook, T; Davidson, A; Dodd, JW; Gormley M; Gregson, F; Hamilton, F; Maskell, N; Morley, A; Murray, J; Keller, J; Pickering, A.E; Reid, J; Sheikh, S; Shrimpton, A; White, C. <u>Author contributions:</u> JD, NM, FH, FG, DA, GN and JB designed the experiments. CR performed the lung physiology testing. SS and FG analysed the data, with BB and JR providing supervisory support and analysis. GN performed supplementary experiments.

## Abstract

**Introduction:** Lung function tests are fundamental diagnostic and monitoring tools for patients with respiratory symptoms. There is significant uncertainty around whether potentially infectious aerosol is produced during different lung function testing modalities; and limited data on possible mitigation strategies to reduce risk to staff and limit fallow time.

**Methods:** Healthy volunteers were recruited in an ultraclean, laminar flow theatre and had standardised spirometry as per ERS/ATS guidance, as well as peak flow measurement and FENO assessment of airway inflammation. Aerosol emission was sampled minimum once each second using both an Aerodynamic Particle Sizer (APS) and Optical Particle Sizer (OPS), and compared to breathing, speaking and coughing. Mitigation strategies such as a peak flow viral filter and a CPET facemask (to mitigate induced coughing) were tested.

**Results:** 33 healthy volunteers were recruited. Aerosol emission was highest in cough (1.61 particles/cm^3^/sample), followed by unfiltered peak flow (0.76 particles/cm^3^/sample). Filtered spirometry produced lower peak aerosol emission (0.11 particles/ cm^3^/sample) than that of a voluntary cough, and addition of a viral filter to the mouthpiece reduced peak flow aerosol emission to similar levels. The filter made little difference to recorded FEV peak flow values. Peak aerosol FENO measurement produced negligible aerosol. Reusable CPET masks with filter reduced aerosol emission when breathing, speaking, and coughing significantly.

**Conclusions:** Compared to voluntary coughing, all lung function testing produced fewer aerosol particles. Filtered spirometry produces lower peak aerosol emission than peak voluntary coughing, and should not be deemed an aerosol generating procedure. The use of viral filters reduces aerosol emission in peak flow by > 10 times, and has little impact on recorded peak flow values. CPET masks are a useful option to reduce aerosol emission from induced coughing while performing spirometry.

## Introduction

The coronavirus disease 2019 (COVID-19) pandemic has had a considerable impact on non-COVID-19 medical services. During the peaks of the pandemic, due to concerns about viral transmission the majority of respiratory diagnostic services were cancelled or delayed (1, 2). As infection rates continue to fluctuate and new severe acute respiratory syndrome coronavirus 2 (SARS-CoV-2) variants emerge (3) there is an urgent need to mitigate risks to patients and staff from transmission and to re start these vital diagnostics services.

SARS-CoV-2 transmission is believed to occur through four modes: direct contact, indirect contact via contaminated objects, droplet production and bioaerosol formation (4). Droplets generally refer to particles >5µm that rapidly fall from the air, whereas bioaerosols are pathogen-containing particles <5µm that can remain suspended in the air for hours. Simple infection control procedures such as handwashing, social distancing and wearing of fluid resistant surgical masks (FRSM) will protect against the first three modes of transmission (5). Such measures are not always effective against bioaerosols. The World Health Organisation defines an aerosol-generating procedure (AGP) as any ‘procedure on a patient that can induce the production of aerosols of various sizes’ (6). Health bodies mandate that additional infection control measures are employed when AGPs are performed (6, 7), these include the use of FFP3/N95 facemasks, patient segregation and enhanced ventilation as measured by the number of air changes per hour (ACH) which dictates the ‘fallow’ time between procedures. These all have time and cost implications, with the alternative approach being to avoid AGPs unless they are a necessity.

Respiratory diagnostics including spirometry, peak expiratory flow rate (PEFR) and fractional exhaled nitric oxide (FENO) measurements which are commonly used in both community and hospital setting. Lung function tests aide diagnosis, monitoring and management for a range of respiratory conditions, as well as to assess fitness for anaesthesia or invasive procedures such as lung biopsy or targeted anti-cancer therapies. Forced expiratory manoeuvres during spirometry and peak flow might promote aerosol production due to shear stress produced between high-speed airflow with the mucus-lined respiratory tract (8) and through the reopening of terminal bronchioles after exhalation to residual volume (9, 10).

A rapid systematic review identified seven sources that classified the AGP status of lung function test (11). Four of these defined lung function tests as an AGP, two defined them as a possible AGP and one source stated that were not an AGP. All cited sources were guidelines, and none referred to primary experimental data that underlay these suppositions. Current advice from expert groups and guidance bodies is contradictory. For example, a working group for the European Respiratory Society suggested full personal protective equipment should be worn for all lung function testing and that they should only be performed when absolutely necessary (1). In contrast, Public Health England do not deem lung function test to be an AGP (12). Finally, the Association for Respiratory Technology and Physiotherapy (ARTP) suggest a more nuanced approach depending on local risk mitigation, ventilation, and local case levels (13).

Primary data on the relative and absolute risk of lung function testing is therefore urgently needed. In this study, we measured aerosol emission from healthy volunteers undergoing spirometry in an ultra-clean, laminar flow theatre with very low background aerosol emission, and compared with patients breathing, speaking, and coughing. We also assess the impact of aerosol mitigation measures including standard mouthpiece viral filters (for spirometry and peak flow) and a proof of concept mitigation measure to reduce the impact of procedure provoked cough aerosol emission. This was done by testing a small group of subjects wearing a close fitting standard cardiopulmonary exercise (CPET) face mask with viral filter which could be adopted prior to entry to the room where lung function testing takes place.

## Methods

### Ethics

This study was performed as part of the wider AERATOR study to assess the risk of aerosolised transmission of SARS-CoV-2 in healthcare settings. Ethical approval was given by the North West Research Ethics Committee (Ref: 20/NW/0393, HRA Approved 18/9/20).

### Environmental set up, recruitment, and procedures

Full technical methods are in the supplementary appendix (S1), and have been reported elsewhere.^4^ In brief, healthy volunteers were recruited to allow collection of aerosol emission in ultra-clean, laminar flow operating theatres. Participants underwent a protocolised set of testing (breathing, speaking, and coughing) followed by lung function testing designed to represent standard clinical practice. Time stamps were used to indicate timing and duration of tidal breathing and forced manoeuvres including formal spirometry (as per ATS/ERS guideline(14)), peak flow measurement, and FENO measurement. All testing was performed by an accredited lung function technician using The spirometers systems used single use bacterial viral filters (BVF), peak flow meters were tested both with and without BVF (see supplement). A proof-of-concept test of mitigation strategy to reduce the risk of procedure induced cough aerosolisation was undertaken using a re-usable full face cardiopulmonary exercise facemask with BVF in 3 subjects. The sampling funnel was positioned over the point of greatest exhalation flow.

Measurements of aerosol were taken simultaneously by two separate devices, the Aerodynamic Particle Sizer (APS), and Optical Particle Sizer (OPS), both manufactured by TSI Ltd), via a 3D printed funnel and through 0.45m of non-conductive tubing. Measurements of aerosol were taken at the exit port of spirometer (for spirometry), the end of the peak flow device (for peak flow), and at various points around the device (for FENO), within 15cm of the source of aerosol..

For brevity, we report only the APS figures as we have previously shown correlation between the devices to be excellent (r> 0.95) (15).

## Results

### Demographics

33 healthy volunteers were recruited; with demographics and lung function results in Table 1. They were young (median age 32), of normal weight (median BMI 23.6) and had normal lung function; as would be expected from this cohort. 16 (48%) were male, with 17 being female (52%).

**Table 1:**
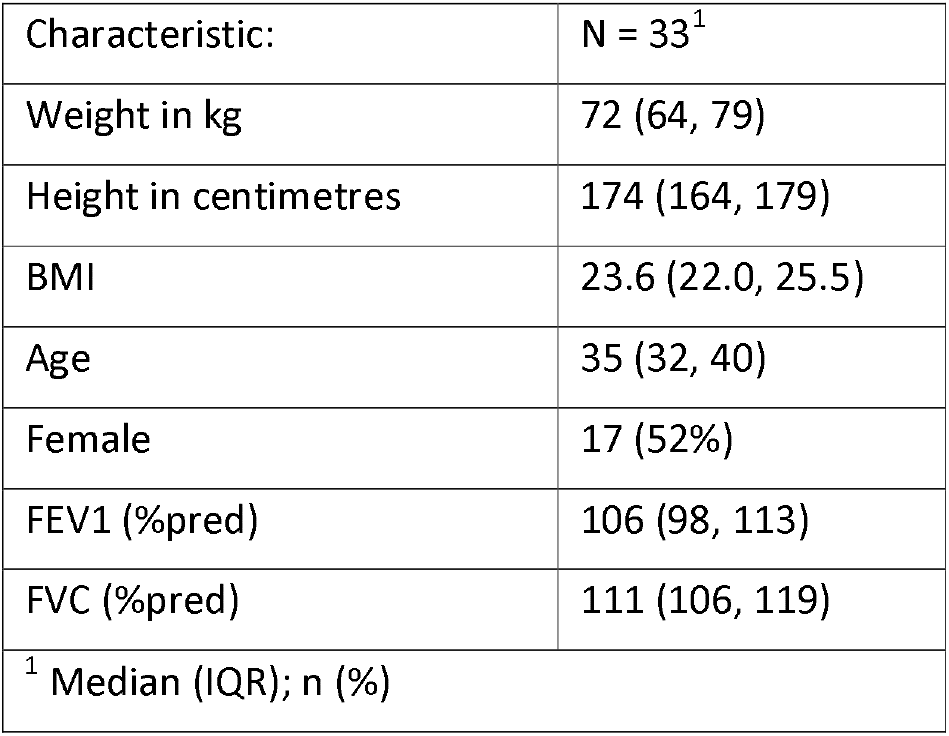
Demographics of the study cohort

|  |  |
| --- | --- |
| Characteristic: | N = 33<sup>1</sup> |
| Weight in kg | 72 (64, 79) |
| Height in centimetres | 174 (164, 179) |
| BMI | 23.6 (22.0, 25.5) |
| Age | 35 (32, 40) |
| Female | 17 (52%) |
| FEV1 (%pred) | 106 (98, 113) |
| FVC (%pred) | 111 (106, 119) |
| <sup>1</sup> Median (IQR); n (%) |  |

### Spirometry

Table 2 describes the geometric mean and standard deviation for FEV1, peak flow, and FENO, in comparison with speaking, breathing and coughing, with Figure 1 describing this on a boxplot. As all of those activities apart from breathing and speaking are sporadic activities, we report the max particles per cm^3^ per sample. Of all the activities, cough produces by far the most, with an average of 1.61 particles/cm^3^/sample, with unfiltered peak flow approximately half (0.76 particles/cm^3^/sample). For brevity, we only discuss the APS results as correlation between the devices is so high.

**Table 2:** Aerosol emission detected from respiratory activities (APS results reported only)

| Activity | Geometric mean particles /cm<sup>3</sup> | Geometric standard deviation particles /cm<sup>3</sup> |
| --- | --- | --- |
| Background | <0.001 | n/a |
| Breathing | 0.04 | 3.37 |
| Speaking | 0.10 | 1.89 |
| Voluntary cough* | 1.61 | 43.06 |
| Peak flow (w/o filter)* | 0.76 | 3.21 |
| Peak flow (w/ filter)* | 0.09 | 1.59 |
| FEV<sub>1</sub> * | 0.11 | 2.10 |
| FENO* | <0.001 | n/a |
| CPET (breathing) | 0.003 | n/a |
| CPET (speaking) | <0.001 | n/a |
| CPET (cough) * | 0.16 | 8.65 |
| As described in the methods; *sporadic activities are calculated from the *peak* aerosol concentration as indicated; whereas continuous activities are calculated from the *average* aerosol concentration.<br>All measurements are APS |  |  |

**Figure 1:**
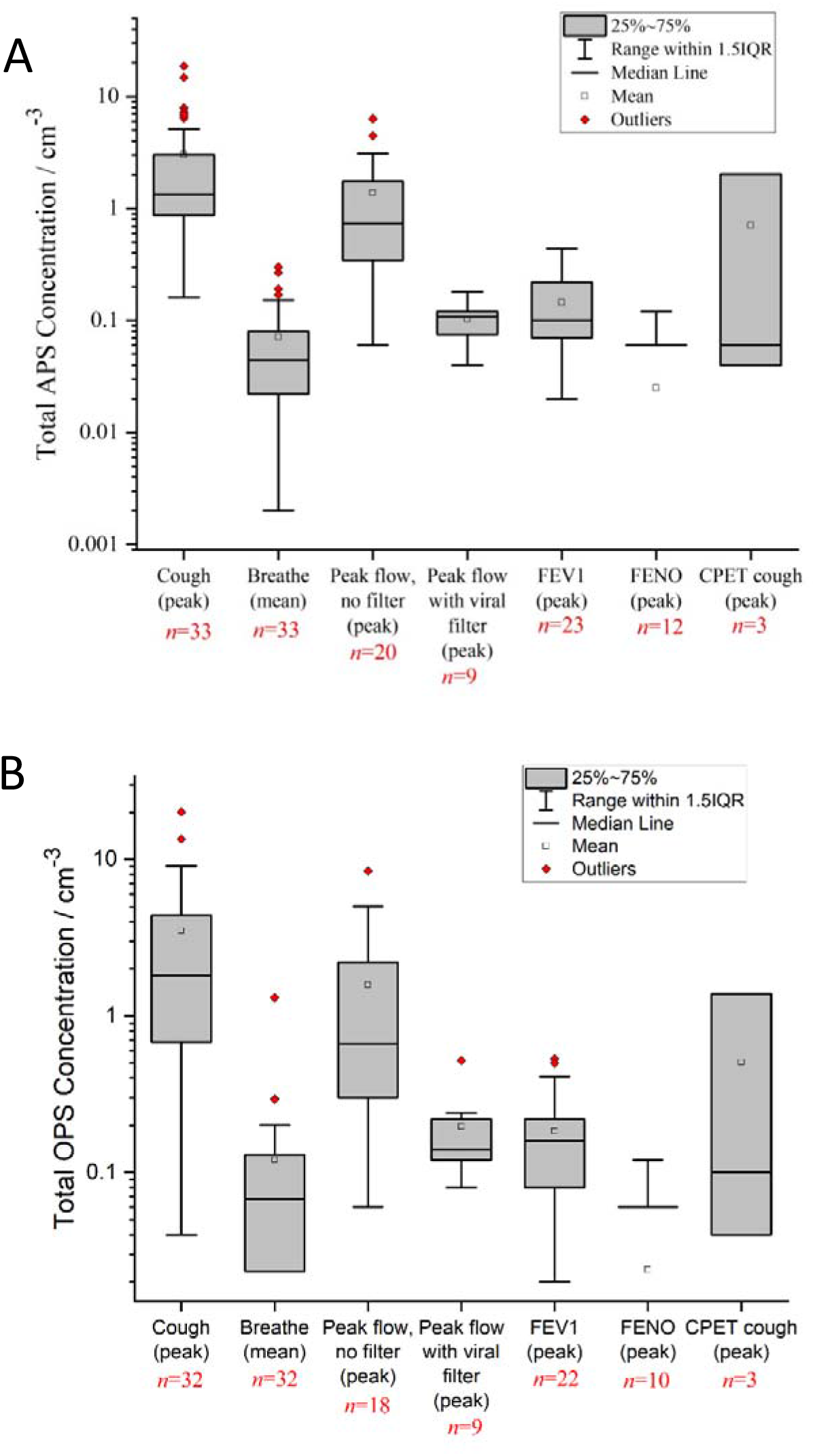
Concentrations of aerosol detected from spirometric activities (a: APS, b: OPS)

Usage of a filter on the peak flow device reduced aerosol emission to a peak emission comparable with the average emission produced while speaking (0.09 particles /cm^3^/sample vs 0.104 particles/cm^3^/ sample). Performing an FEV1 had similar peak aerosol emission (0.11 particles /cm^3^/ sample).

The FENO device does not have a clear exhalation port and the manufacturer does not make clear where exhaled breath leaves the device. We interrogated the device and measured aerosol at all possible exhalation ports (see image in supplementary appendix). Regardless, we did not find any significant aerosol emission from the FENO device.

Usage of a CPET mask to reduce aerosol emission was tested on 3 participants, with large reductions in aerosol emission during breathing, speaking and coughing (results in Table 2). This mask can be worn before, during, and after spirometry and may be a potential mitigation technique as induced coughing is common during lung function.

In summary, formal lung function testing using a spirometer with a filter did not generate significant aerosol in comparison to a cough; with peak levels similar to average emissions during speaking.

### Peak flow with a filter

To ensure peak flow results were reliable with a filter, we compared peak flow readings both with and without the protective filter for 9 participants. This is shown in figure 2, showing strong correlation with a R of 0.966. Therefore, although there may be a slight reduction in recorded FEV1 with the peak flow monitor when used with a filter, this is likely clinically insignificant.

**Figure 2:**
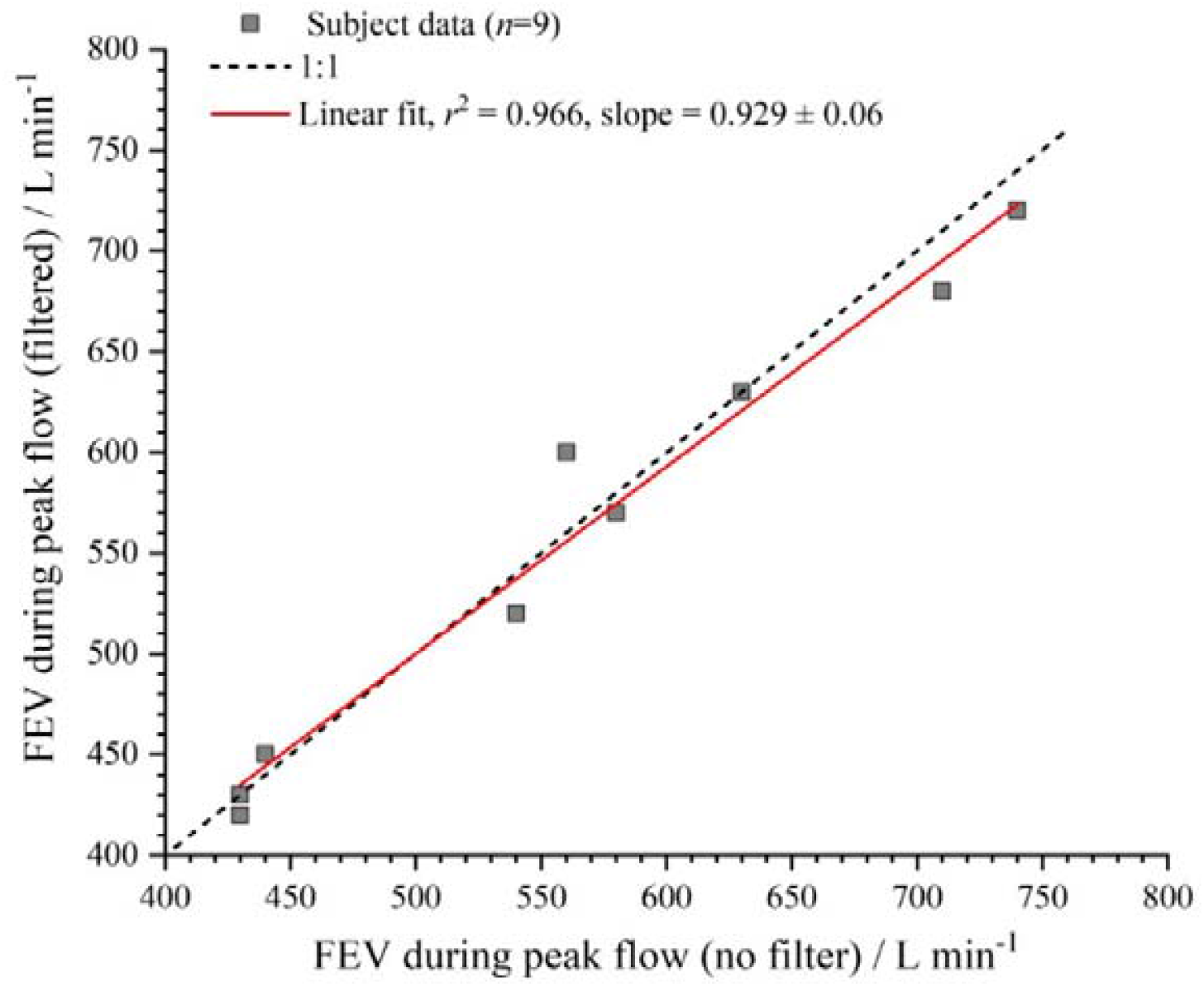
Correlation between unfiltered and filtered peak flow

### Aerosol emission and relation to spirometric function

Finally, we investigated the relationship between predicted and total FEV1 and FVC with aerosol emission during speaking, breathing and coughing, and found no clear relationship (Figure 3, r <0.4 for all comparisons.)

**Figure 3:**
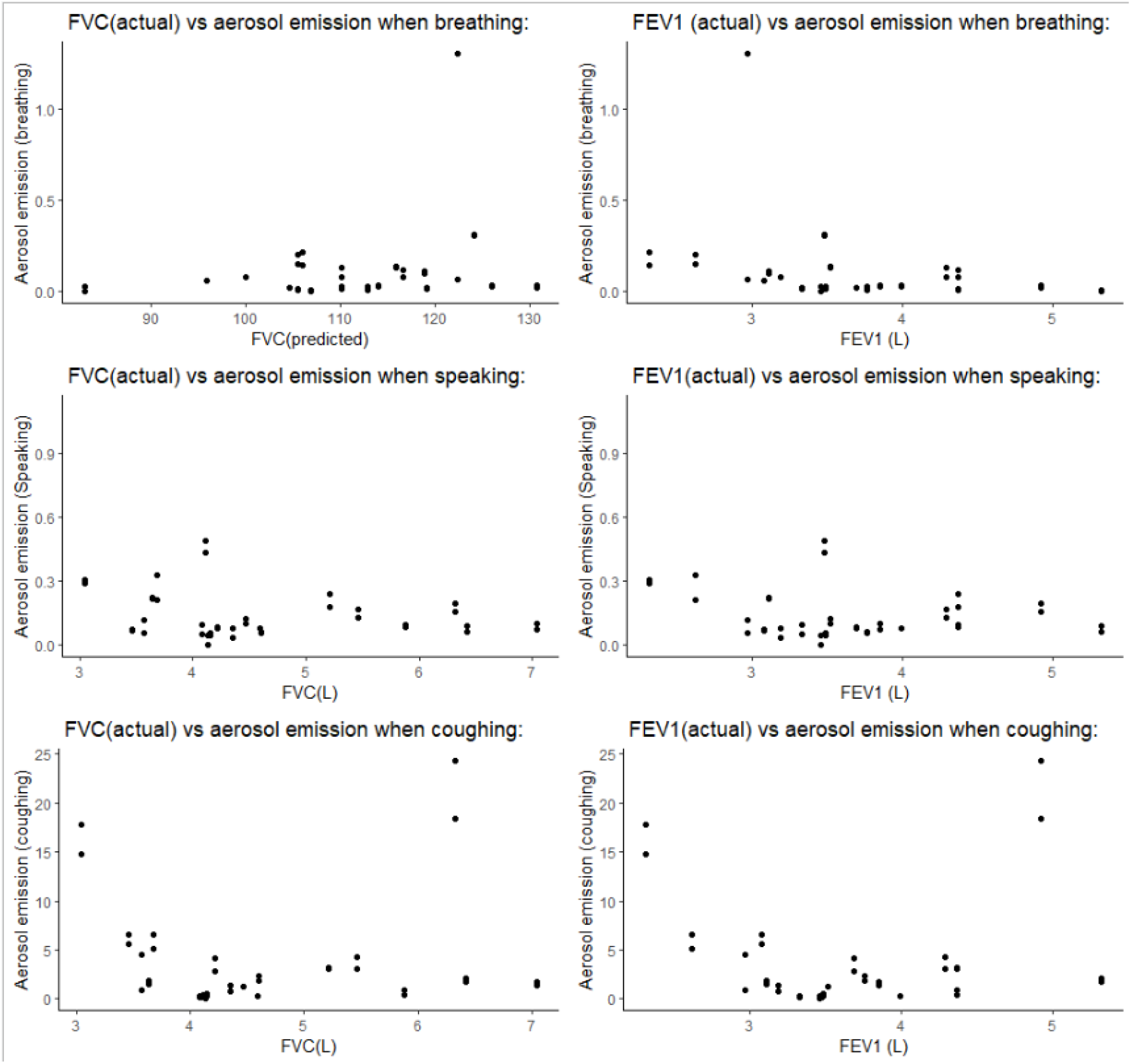
FVC and FEV1 compared to aerosol emission when breathing, speaking and coughing.

## Discussion

Pulmonary function tests including spirometry, peak flow and exhaled nitric oxide are a fundamental component of the evaluation of patients with pulmonary diseases. They require subjects to blow into a tube which measures volume and flow of expired air, in the case of spirometry and peak flow these are forced manoeuvres. In the absence of robust experimental data, the precautionary principle necessitated policy makers to designate them as aerosol generating procedures mandating considerable adaptations to be put in place due to COVID-19. These included pre-clinic COVID-19 screening, donning full PPE, room cleaning and fallow time to allow multiple air changes between patients. This has reduced capacity resulting in significant pressures on both primary and secondary care to provide equitable, safe and timely access to respiratory diagnostics.

This study provides much needed high quality experimental data characterising aerosol emissions during clinical standard respiratory clinical diagnostic tests. The finding that spirometry, peak flow and exhaled nitric oxide do not generate clinically significant aerosol (in comparison to cough) is reassuring and will inform service providers, commissioners and policy and guideline makers and should support plans to safely re open services and address the back log in respiratory diagnostics and monitoring.

Peak flow testing without a filter does produce aerosol (although less than in coughing) however, the addition of a filter reduces this aerosol to negligible levels and renders the procedure non aerosol generating. Together this points to the high effectiveness of stand CE marked viral filters in reducing aerosol emission. The study also identifies a potential effective mitigation measure (re-usable filtered CPET mask) for aerosols generated by subjects who cough as a result of the procedure. We believe that this may allow for a significant increased diagnostic capacity through reduced need for air room changes between subjects.

### Strengths and weaknesses

This study has many strengths. Firstly, we performed spirometry as per standard UK practice in an ultraclean, laminar flow theatre with very little background aerosol concentration. This allows us to make confident conclusions about the source and level of aerosol produced. Secondly, we were able to test mitigation strategies allowing for simple practical guidance on risks. Thirdly, we used two simultaneous measurement techniques (OPS and APS), that ensured robustness of measured results. Finally, we were able to compare to a known reference (cough), as a comparison for a procedure that does generate significant aerosol.

Common to all volunteer studies, we are limited in that our volunteers have largely normal spirometry. Reassuringly, however, we found no relationship between aerosol emission and FEV1 or FVC, and there is no evidence available from the current literature to suggest that patients with lung disease are more likely to generate aerosol. The AERATOR study will include further aerosol emission data from patients with underlying lung disease and increase the number of healthy volunteers wearing CPET mask in the near future which will improve sample size and address this question. Also, none of the volunteers were known to be suffering from viral infections which have been shown to increase aerosol production (16, 17). However, while the available aerosol data from patients with active COVID-19 is limited they do suggest comparable emissions to healthy volunteers (15). Finally, as activities such as coughing or peak flow are forceful and short lived, these are observed as a rapid rise in the reported number of aerosol concentration, then a decay away over a few sample measurements (typically equivalent to 10-15 s for a cough) as the aerosol dissipates from the sampling funnel and is diluted by the clean room air. This means the comparison between these forceful events and more continuous events (such as breathing, or speaking) in total aerosol emission is difficult.

### Comparisons with previous literature

There is a lack of high-quality experimental data to reconcile current conflicting guidelines. We identified three recent papers that have attempted to address the issue. Helgeson et al. undertook a small-scale study that identified aerosol generation during spirometry performed by five healthy volunteers (18). It detected aerosol production despite the inclusion of viral filters, though this was not evident when samples 1.5 foot from the exhalation port. It was performed in a room with a good ventilation (12.8 air changes/hr (ACH)). The sampling method used in this study did not detect any increase in aerosol production with speaking bringing into questions the sensitivity of the methodology. Furthermore, data was discarded if a participant coughed. We consider this an important and relevant reference for other tested AGPs. Lastly, their small sample size limits generalisability.

The second study (19) included 33 healthy volunteers who performed different respiratory manoeuvres outside of ultra clean, environment. Like our data they found coughing was the highest aerosol-producing manoeuvre and interestingly they found that increased expiratory flow rates did not increase aerosol production. The study was performed without viral filters and thus does not provide data relevant to this simple mitigation factor. Furthermore, the authors acknowledge that their PFTs did not meet ERS/ATS spirometry criteria, and 22 of the volunteers performed nothing more than tidal breathing.

The third paper (20) analysed aerosol presence whilst a variety of pulmonary function tests (PFTs) were performed by 28 patients with viral filters in rooms with three different ACH rates. Just one measure of aerosol (OPS) was performed during PFTs and for 30-60 minutes following the procedure after the patients and researchers had left the room. The paper concludes that PFTs should be considered an AGP, however on examination of the presented data, during the PFT procedures, aerosol levels remain steady or fall. The peak aerosol production occurs when the patients move away from the apparatus and are breathing, speaking and coughing without a mask on. There are no data presented to show baseline aerosol levels with patients in the room without masks on, and thus we can only conclude that the data shows the relatively low level of aerosol production during PFTs and re-emphasises the importance of mask-wearing and social distancing during normal interactions. It also highlights the potential benefits of the use of measures such as a CPET mask in preventing aerosol production in the time between procedural manoeuvres.

### Significance of this study

This study has demonstrated that peak flow measurement and spirometry performed with viral filters, and standard FENO do not promote significant aerosol production. It supports their removal from the designated list of AGPs. This should allow community and hospital-based lung physiology teams to increased capacity for these essential respiratory diagnostics by reviewing the necessity for restrictive measures such as community PCR testing, patient self-isolation prior to attendance, the use of ‘full PPE’ during procedures, and prolonged fallow periods during which time further procedures cannot be performed.

## Conclusions

Spirometry (performed standard filter) and FENO do not generate clinically significant aerosol. Peak flow does generate aerosol (around half of that of a cough), although a viral filter reduces this to levels similar to speaking. Re-usable CPET masks with filters applied to subjects prior to testing is a potential solution to mitigate aerosol emission from induced coughing before and after the procedure.

## Data Availability

Raw data will be available with the full paper at the Bristol University Data Repository.

http://www.data.bris.ac.uk

## References

1. McGowan A, Sylvester K, Burgos F, Boros P, De Jongh F, Kendrick A, et al. Recommendation from ERS Group 9.1 (Respiratory function technologists /Scientists) Lung function testing during COVID-19 pandemic and beyond. 2020 [Available from: https://www.artp.org.uk/write/MediaUploads/Standards/COVID19/ERS_9.1_Statement_on_lung_function_during_COVID-19_Version_1.0.pdf.

2. ARTP/BTS. Respiratory Function Testing During Endemic COVID-19 2020 [updated 26/05/2020. Available from: https://www.artp.org.uk/write/MediaUploads/Standards/COVID19/Respiratory_Function_Testing_During_Endemic_COVID_V1.5.pdf.

3. Phillips N. The coronavirus is here to stay - here’s what that means. Nature. 2021;590(7846):382–4.

4. Dhand R, Li J. Coughs and Sneezes: Their Role in Transmission of Respiratory Viral Infections, Including SARS-CoV-2. American Journal of Respiratory and Critical Care Medicine. 2020;202(5):651–9.

5. PHE. COVID-19: personal protective equipment use for non-aerosol generating procedures. 2020 [updated 21/10/2020. Available from: https://www.gov.uk/government/publications/covid-19-personal-protective-equipment-use-for-non-aerosol-generating-procedures.

6. WHO. Infection Prevention and Control of Epidemic- and Pandemic-Prone Acute Respiratory Infections in Health Care. Geneva: World Health Organization; 2014 [Available from: https://apps.who.int/iris/handle/10665/112656.

7. PHE. COVID-19: personal protective equipment use for aerosol generating procedures. 2020 [updated 21/08/2020. Available from: https://www.gov.uk/government/publications/covid-19-personal-protective-equipment-use-for-aerosol-generating-procedures.

8. Wei J, Li Y. Airborne spread of infectious agents in the indoor environment. Am J Infect Control. 2016;44(9 Suppl):S102–8.

9. Almstrand A-C, Ljungström E, Lausmaa J, Bake B, Sjövall P, Olin A-C. Airway Monitoring by Collection and Mass Spectrometric Analysis of Exhaled Particles. Analytical Chemistry. 2009;81(2):662–8.

10. Johnson GR, Morawska L. The mechanism of breath aerosol formation. J Aerosol Med Pulm Drug Deliv. 2009;22(3):229–37.

11. Jackson T, Deibert D, Wyatt G, Durand-Moreau Q, Adisesh A, Khunti K, et al. Classification of aerosol-generating procedures: a rapid systematic review. BMJ Open Respir Res. 2020;7(1).

12. PHE. COVID-19 infection prevention and control guidance: aerosol generating procedures. 2020 [updated 21/01/2021. Available from: https://www.gov.uk/government/publications/wuhan-novel-coronavirus-infection-prevention-and-control/covid-19-infection-prevention-and-control-guidance-aerosol-generating-procedures.

13. Group AC-. Association for Respiratory Technology and Physiology (ARTP) – Guidelines for recommencing physiological services during the Coronavirus Disease 2019 (COVID-19) endemic phase: version 5.1. 2020 [updated 24/08/2020. Available from: https://www.artp.org.uk/write/MediaUploads/Standards/COVID19/ARTP_COVID-19_endemic__guidance_Vers_5.6_final.pdf.

14. Graham BL, Steenbruggen I, Miller MR, Barjaktarevic IZ, Cooper BG, Hall GL, et al. Standardization of Spirometry 2019 Update. An Official American Thoracic Society and European Respiratory Society Technical Statement. American Journal of Respiratory and Critical Care Medicine. 2019;200(8):e70–e88.

15. Hamilton F, Gregson F, Arnold D, Sheikh S, Ward K, Brown J, et al. Aerosol emission from the respiratory tract: an analysis of relative risks from oxygen delivery systems. medRxiv. 2021:2021.01.29.21250552.

16. Lindsley WG, Pearce TA, Hudnall JB, Davis KA, Davis SM, Fisher MA, et al. Quantity and size distribution of cough-generated aerosol particles produced by influenza patients during and after illness. J Occup Environ Hyg. 2012;9(7):443–9.

17. Hersen G, Moularat S, Robine E, Géhin E, Corbet S, Vabret A, et al. Impact of Health on Particle Size of Exhaled Respiratory Aerosols: Case-control Study. Clean (Weinh). 2008;36(7):572–7.

18. Helgeson SA, Lim KG, Lee AS, Niven AS, Patel NM. Aerosol Generation during Spirometry. Annals of the American Thoracic Society. 2020;17(12):1637–9.

19. Greening NJ, Larsson P, Ljungström E, Siddiqui S, Olin A-C. Small droplet emission in exhaled breath during different breathing manoeuvres: Implications for clinical lung function testing during COVID-19. Allergy.

20. Li J, Jing G, Fink JB, Porszasz J, Moran EM, Kiourkas RD, et al. Airborne Particulate Concentrations During and After Pulmonary Function Testing. Chest. 2020.

