## Supplement S1 for "Risk and mitigation of aerosolisation from lung function testing: results from the AERATOR study" (from another version of this preprint: version 1, 2021-03-08)

Investigation of respiratory disease: risk of aerosolisation from spirometry, peak flow, and other associated tests.

*These authors contributed equally

#These authors contributed equally

Corresponding author:

ORCID 0000-00034805-5759

*Ethics:*

This study was performed as part of the wider AERATOR study to assess the risk of aerosolised transmission of SARS-CoV-2 in healthcare settings. Ethical approval was given by the North West Research Ethics Committee (Ref: 20/NW/0393, HRA Approved 18/9/20).

*Aerosol measurement*

Aerosol measurements were recorded using two devices simultaneously: an Optical Particle Sizer (OPS) and Aerodynamic Particle Sizer (APS). Technical specifications were detailed in a previous publication from our group but are replicated here.[1,2]

The APS (TSI Incorporated, model 3321, Shoreview, NM, USA) measures aerosol at a sampling flow rate of 1 L min^-1^ with accompanying sheath flow of 4 L min^-1^. The APS reports the aerodynamic size of particles in an aerosol plume, size-resolving aerosol number concentration into 52 size bins ranging from 0.5 µm to 20 µm in diameter with a time integration of 1 s. The size bins are equally spaced in log(diameter) space, apart from the smallest size bin (0.5 - 0.523 µm).

The OPS (TSI Incorporated, model 3330, Shoreview, NM, USA) samples air at 1 L min^−1^ and detects particles by laser optical scattering. The OPS reports the particle number concentration and optical size distribution within the diameter range 300 nm to 10 µm with a time resolution of 1 second. The OPS is widely used for aerosol studies from laboratories / clean rooms to more demanding outdoor environments. It is calibrated by the manufacturer using polystyrene latex spheres and its performance conforms to the ISO standard 21501‐4:2018. The reported optical size of the particles is based on an assumed refractive index of pure water at 600 nm wavelength (1.333).

Both the APS and OPS were connected to the same sampling funnel, which was 3D printed (RAISE3D Pro2 Printer, 3DGBIRE, Chorley, UK) from PLA with a maximum diameter of 150 mm, cone height of 90 mm with a 10‐mm exit port. Two conductive silicone sampling tubes of 0.3 m length and internal diameter 4.8 mm (3001788, TSI) were connected to the neck of the sampling funnel, with one connected to the APS and the other to the OPS.

For baseline measurements and HFNO, the funnel was placed such that the top of the funnel cone was 1 cm from the participant’s forehead, and the sampling apex of the funnel was 10 cm from the participant’s mouth. For CPAP the sampling apex of the funnel was 10 cm from the exit port or area of greatest facemask leak. Supplement 1 includes some images from the set up to aid visualisation.

*Environmental set up and patient recruitment*

Healthy volunteers were recruited in an ultra-clean laminar flow operating theatre (EXFLOW 32, Howarth Air Technology, Farnworth, UK) with high efficiency particulate air (HEPA) filtration and an air supply rate of 1200 m^3^/s. This ventilation system has a canopy ‘clean zone’ where surgical procedures are performed; the air circulation velocity is 0.2 m.s^−1^ at 1 m above the floor below the canopy and produces 500–650 air changes per hour. All aerosol recordings were performed under the canopy, and the background aerosol concentration was sampled prior to each measurement for a mean sampling duration of 43 s.

*Statistical analysis*

Aerosol generation differs greatly between people, with an approximate log-normal distribution in number concentration.[3,4] As such, our analysis focussed on comparing the relative aerosol number concentrations from different procedures performed by each individual. We report the number concentration, an intensive property that does not depend on scale (i.e. is independent of the time or volume sampled) as reported by the instruments measured over a sample period, selected to be 1 s. We have reported one of two parameters for each activity: either the peak particle number concentration reported across the full number of samples of the measurement for single, forced exhalations such as coughing (cm^-3^); or, the mean particle number concentration reported as the average across all samples for continuous activities such as breathing or speaking (cm^-3^). We then visualised size distributions of aerosol emission across the volunteers and compared aerosol emission across activities. Data analysis was performed by collating raw data of sampled aerosol concentration output by the APS and OPS instruments using Aerosol Instrument Manager 9.0 (TSI Incorporated, Shoreview, NM, USA) and Microsoft Excel. A custom-written software in LabVIEW (National Instruments, Texas, USA) was used to automate the analysis process for increased efficiency.

*Images*

**DUE TO MEDRXIV POLICY, IMAGES CONTAINING HUMAN SUBJECTS ARE NOT PERMITTED. IN THE SUBSEQUENT PUBLICATION IMAGES WITH HUMANS IN WILL BE INCLUDED. PLEASE CONTACT THE CORRESPONDING AUTHOR IF IMAGES ARE REQUIRED BEFORE THEN.**

Figure S1 : Peak flow device without filter

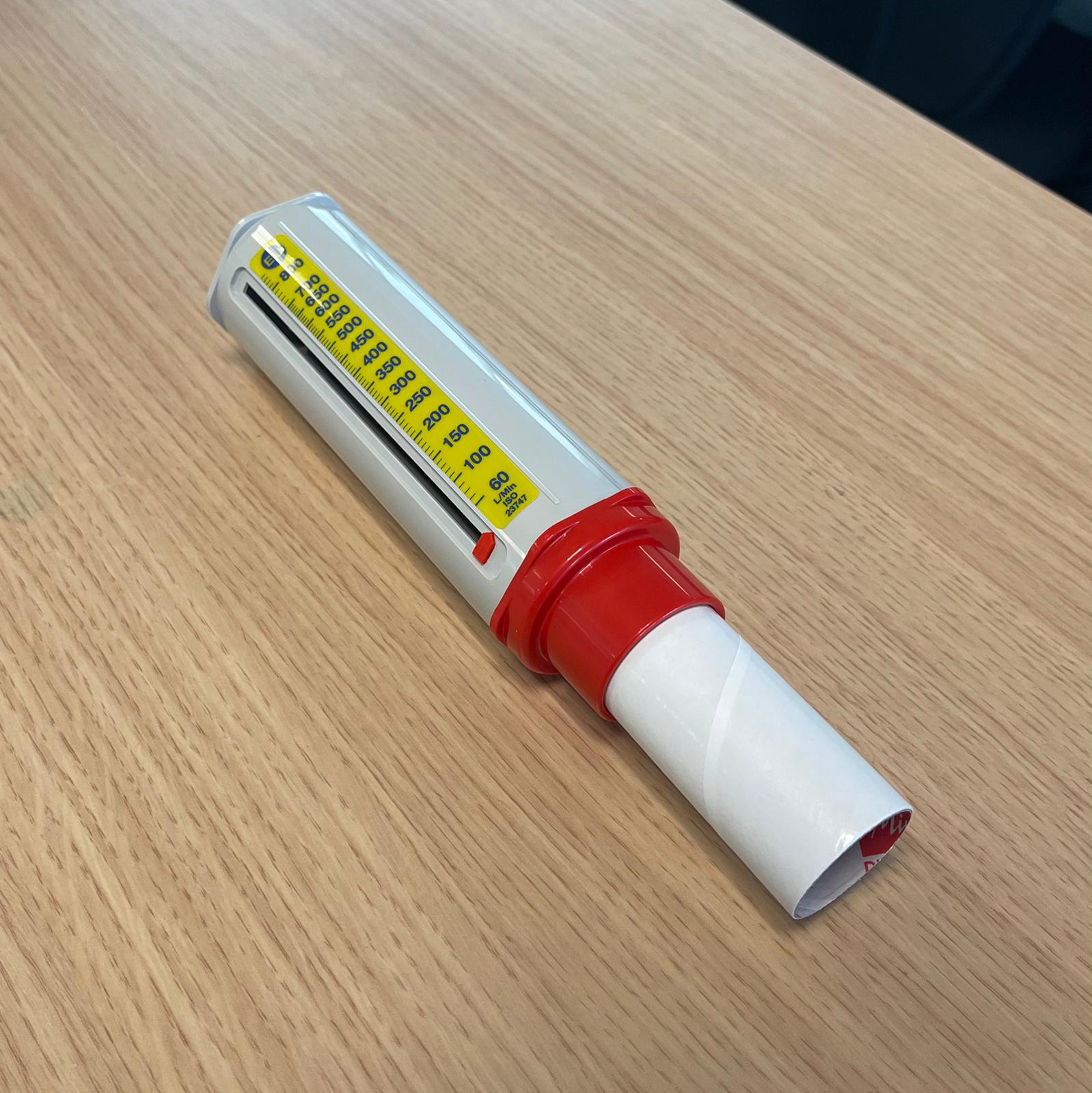

Figure S2: Peak flow device with filter

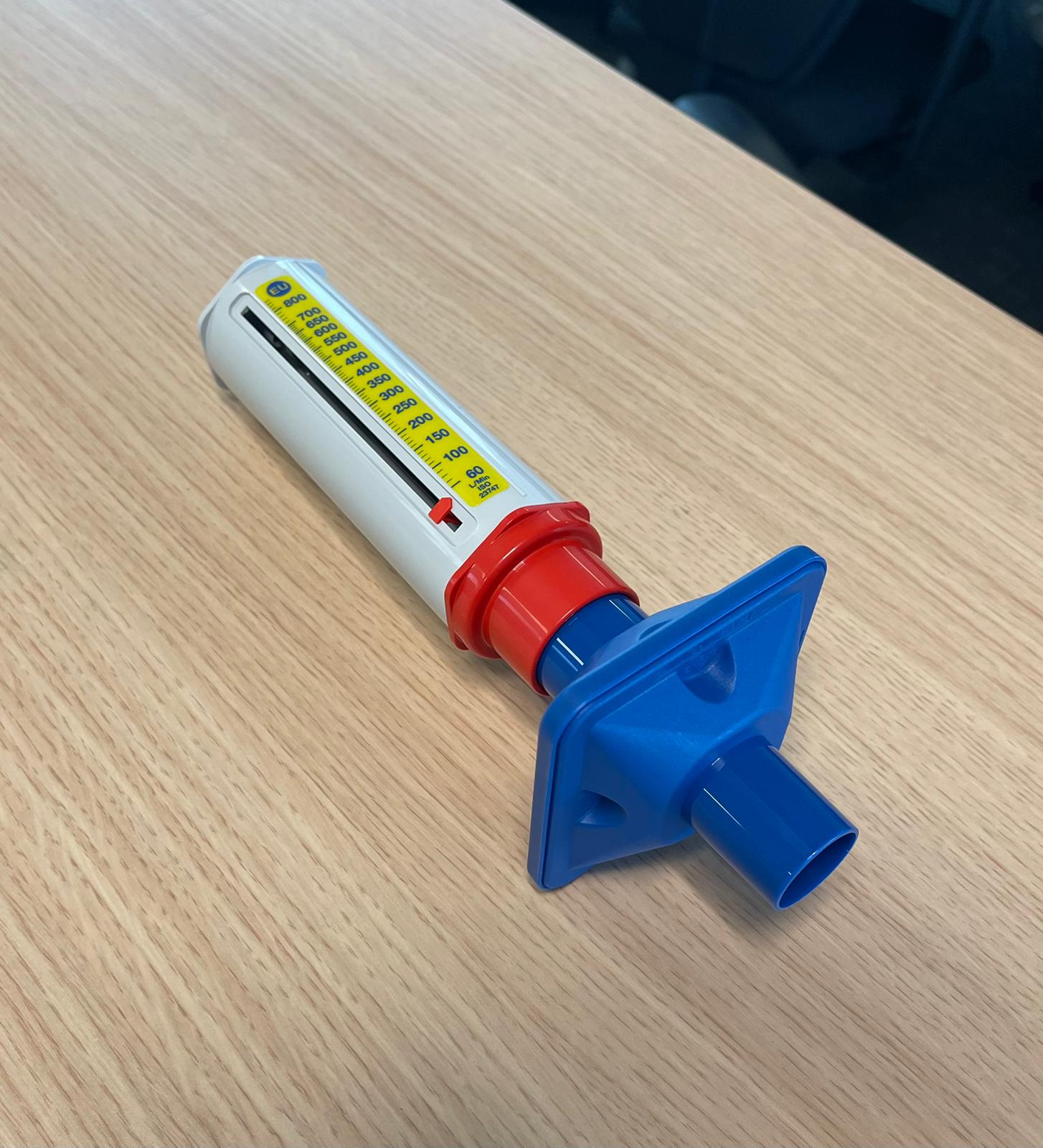

Figure S3: CPET mask

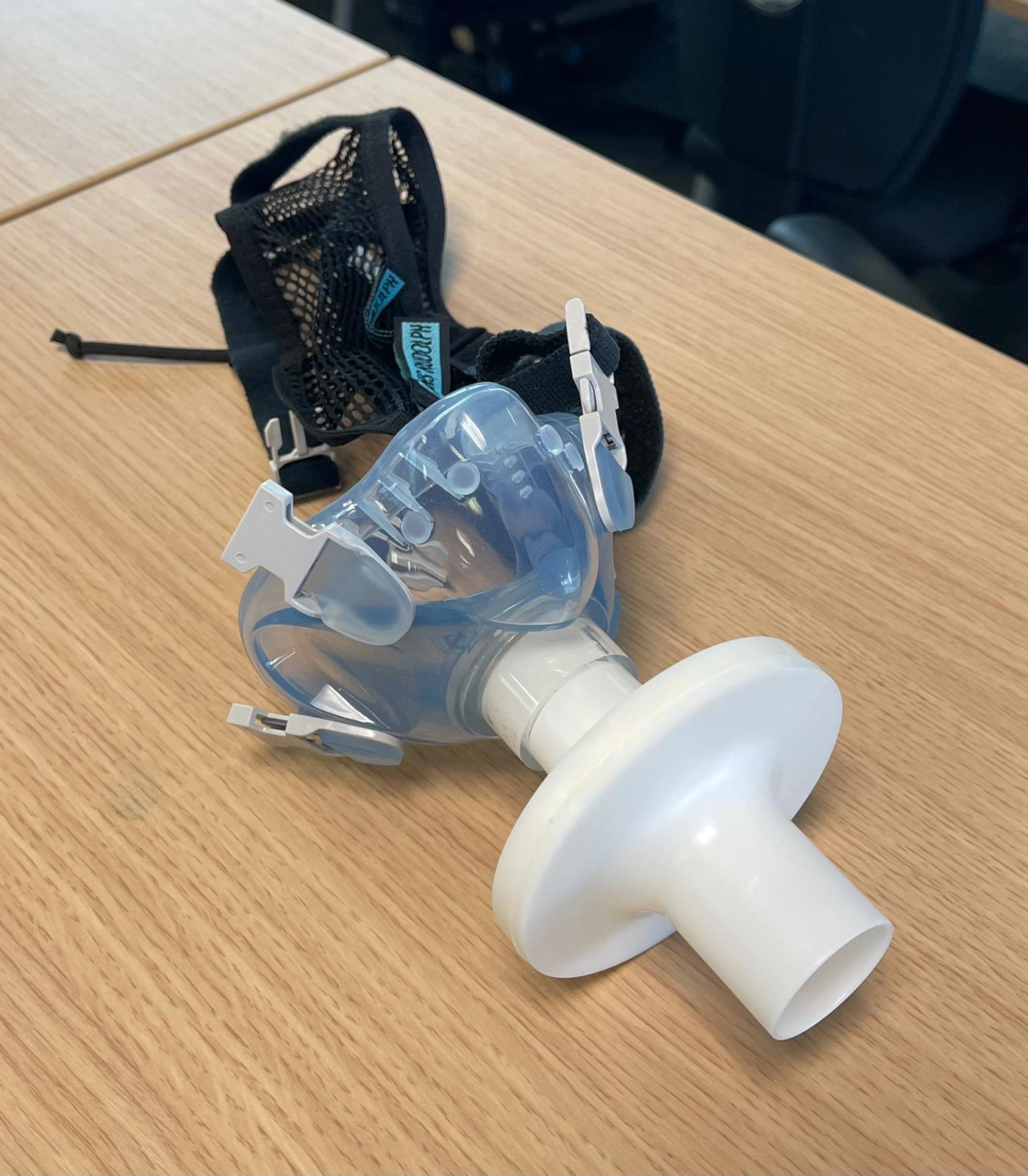
